# The Potential Role of the Regional Skull Conditions in Predicting the Efficacy of Transcranial Magnetic Resonance-guided Focused Ultrasound in Patient with Low Skull Density Ratio

**DOI:** 10.1101/2025.03.12.25323386

**Authors:** Makoto Kadowaki, Kenji Sugiyama, Takao Nozaki, Akira Okazaki, Muneaki Hashimoto, Tomohiro Yamasaki, Yoshinobu Kamio, Mikihiro Shimizu, Namba Hiroki, Kazuhiko Kurozumi

**Author notes:** Corresponding Author’s name and current institution: Takao Nozaki, Department of Neurosurgery, Hamamatsu University School of Medicine, Hamamatsu, Shizuoka, Japan, Corresponding Author’s. Previous Presentations: The 20th Biennial Meeting of the World Society for Stereotactic & Functional Neurosurgery, 2024/09/04, Chicago, USA, e-Poster Presentation.

## Abstract

**Objective:** The therapeutic effect of magnetic resonance-guided focused ultrasound is limited to patients with a low skull density ratio (SDR). We explored the skull conditions associated with successful treatment among low-SDR patients, and, to compensate for the small sample size, performed analyses using all cases irrespective of SDR. This is the first report to examine the significance of regional skull conditions.

**Methods:** We retrospectively analyzed 171 consecutive cases. Descriptive statistics for the entire skull, and averages for 10 regions, were obtained for variables including the SDR, skull thickness, and ultrasound incident angle (IA; smaller = more vertical). The 1,024 ultrasonic transducer elements were divided into 10 regions predefined by ExAblate4000. Symptoms were evaluated by Clinical Rating Scale for Tremor for essential tremor and Unified Parkinson’s Disease Rating Scale tremor score for Parkinson’s tremor. Successful treatment was defined as <half preoperative symptom score at 6 months postoperatively. First, univariate analysis of cases with SDR < 0.40 was conducted to explore candidates for skull conditions associated with successful treatment. Subsequently, for all cases regardless of SDR, several multiple regression models were built to predict the maximum temperature rise, and their performance was compared.

**Results:** Of the cases, 26 had SDR < 0.40, and 15 were successful. Among the cases with SDR < 0.40, IA of the parietal region on the sonication side and SDR of the bilateral temporal region tended to be smaller in the success group (not statistically significant). The maximum temperature was more accurately predicted when IA of the parietal region on the sonication side was included in the model (Akaike information criterion, 777 [from 757]). Furthermore, replacing SDR with SDR excluding the bilateral temporal region enhanced predictions (Akaike information criterion, 777 [from 767]).

**Conclusions:** Even if SDR is low, treatment success may be more attainable if the IA in the parietal region on the sonication side is smaller, or if the SDR excluding the bilateral temporal regions is large.

## Introduction

The skull density ratio (SDR) is one of the key variables in predicting the therapeutic efficacy of transcranial magnetic resonance-guided focused ultrasound (MRgFUS) lesioning.^1-7^ The US Food and Drug Administration recommends MRgFUS thalamotomy only for patients with an SDR > 0.45 ± 0.05.^8^ When SDR is low, the target site temperature is less likely to rise,^1,3,4^ more energy is required,^2,6,9^ heating efficiency decreases,^10,11^ and adverse events increase.^3^ Recently, however, several retrospective^2-4,7^ and one prospective study^12^ reported successful treatment of low-SDR cases, also noting safety.^3,13^ However, the factors determining success or failure in low-SDR cases remain unexplored.

In general, the mean value across the entire treatment field (SDR_mean_) is used as a representative SDR variable for individual patients. ^1-7^ In addition to SDR_mean_, a thin skull thickness, ^9^ a small incident angle (IA) to the skull (i.e., a more [nearly] vertical angle of incidence),^1,11,14^ and a small skull area^1^ are also expected to be effective in predicting treatment efficacy. This suggests that there may be other skull conditions besides SDR_mean_ that are not well known but can predict treatment effects. A smaller SDR frequency distribution skewness (SDR_skewness_) has been reported to have a greater effect than SDR_mean_, ^5^ and customizing the SDRs independently was effective.^15^ This suggests that there may be other valid ways to calculate SDRs besides the SDR_mean_, possibly indicating that the three-dimensional/spatial distribution of SDRs in the skull may be meaningful. However, no studies have examined the significance of the three-dimensional/spatial distribution of SDRs and other skull conditions in subregions of the overall skull.

Recently, machine learning has been reported to predict energy requirements and the maximum temperatures reached using SDR_mean_ and SDR values for individual transducers.^6^ Machine learning is rapidly being adopted in many medical fields, including neurosurgery,^16,17^ and its effectiveness in predicting surgical outcomes has been reported.^17^ Machine learning can process more information than conventional methods and is expected to provide more accurate diagnoses. However, machine learning’s predictive models are difficult to understand in terms of their internal operation, and they do not diminish the significance of conventional predictive models in terms of transparency, explainability, reliability, and simplicity.^18,19^

On the basis of the outcomes of low-SDR cases at our institution, we used conventional statistical methods to explore the cranial conditions associated with successful treatment of patients with a low SDR. In addition to previously reported factors, we examined various descriptive statistics and the validity of the distribution in the cranial crown as an explanatory variable. As detailed in the Methods, the entire skull was divided into 10 regions, and the significance of the mean of each region for various skull conditions, including SDR and IA, was examined as a predictor. To address the small sample size, we also conducted a regression analysis using all cases, regardless of SDR.

## Methods

### [Patients]

Patients treated at our institution between April 2020 and September 2023, who were followed up for 6 months after treatment, were included in the study. In particular, written informed consent was obtained for the treatment of patients with an SDR_mean_ of 0.40 or less after thoroughly explaining the risks of treatment failure and complications. Because of the retrospective design, written consent for study participation was waived, and an opt-out approach was applied via the study website.

### [Treatment and Assessment]

The treatment method was detailed in a previous paper.^20^ In summary, sonication was performed with an Exablate 4000 (Insightec, Tirat Carmel, Israel) system, targeting the ventral intermediate nucleus of the thalamus. Target coordinates were initially set on the basis of the atlas and adjusted using magnetic resonance imaging. First, sonication was performed at a low energy dose, and the temperature was increased to approximately 43–47°C. The displacement between the sonication target and temperature rise was then aligned with magnetic resonance thermometry. The energy dose was then increased to 48–50°C to confirm symptom improvement and the absence of side effects. Finally, sonication was performed with a higher energy dose to raise the temperature to 53–60°C, thermally coagulating the target area. Once the intended temperature was reached, magnetic resonance imaging confirmed coagulation foci in the target area. Additional sonication was performed if necessary.

A temperature rise below 53°C was occasionally experienced because of low SDR_mean_. In such cases, possible temperature elevations were repeated. Treatment was considered complete when sufficient symptom improvement and T2 lesion formation were observed. The procedure was abandoned when continuing treatment was deemed difficult because of intraoperative symptoms such as headache, or when the temperature required for thermal coagulation could not be achieved.

The Clinical Rating Scale for Tremor was used to rate essential tremor, and for Parkinson’s disease, scores for tremor were obtained using the Unified Parkinson’s Disease Rating Scale (part II [item 16] and part III [items 20 and 21]). Treatment success was defined as a symptom rating score less than half of the preoperative score at 6 months postoperatively.

### [Skull Condition Variables]

Cranial variables for each of the 1,024 ultrasound elements were obtained from the ExAblate 4000 treatment recordings (SDR, skull thickness, IA). Various descriptive statistics were calculated for all 1,024 elements: mean, maximum, minimum, median, mode, variance, standard deviation, skewness, and kurtosis.

The 1,024 hemispherically arranged ultrasonic transducer elements were then divided into 10 sections/regions (A–J), as classified by default in the ExAblate 4000 system (Figure 1). On the basis of element positions, the central section (A– D) was divided into four equal parts (64 elements each), and the peripheral section (E–J) was divided into left and right parts, and then into three equal sections (frontal, temporal, and occipital) (128 elements each). Data from the right target treatment were mirrored to invert the left and right sides. The mean values of various cranial variables were calculated for each region (A–J).

**Figure 1.**
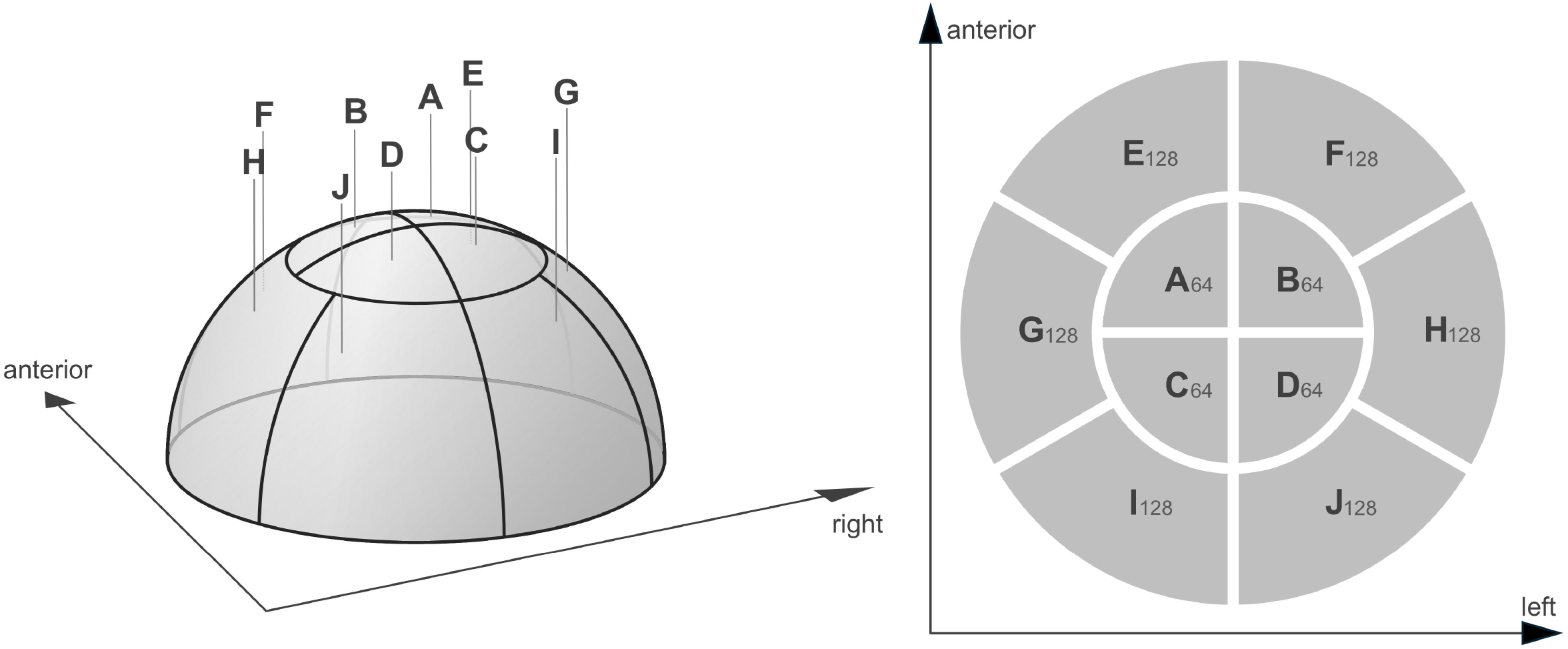
The 1,024 elements are divided into 10 sections. Left, 3D image. Right, unfolded image. Subscripts indicate the number of ultrasonic elements included in each section.

### [Analysis of Cases with SDR_mean_ < 0.40]

We first selected patients with SDR_mean_ < 0.40 from our institution’s treatment database and searched for cranial variables that could influence treatment success or failure. A univariate analysis was performed comparing the various skull condition variables between the success and failure groups, and p-values were obtained. Variables with p < 0.05 were extracted, and their medical significance was assessed.

### [Analysis Using All Cases]

To evaluate the clinical significance of the results in patients with SDR < 0.40, a statistical analysis of all cases, regardless of SDR, was performed. First, the mean or median values of skull condition variables for each of the 10 regions (A– J) were calculated and visualized to detect trends in the overall population, and in the treatment success/failure groups with SDR < 0.40. In all cases, the difference between the mean of each region and the mean of the entire cranium was represented in grayscale. For the success/failure groups of cases with SDR < 0.40, the difference between the median value of each region and the mean value of all cases was shown on a color scale.

Next, the Akaike information criterion^20,21^ (AIC) was calculated by multiple regression analysis, using the candidate variables explored above as explanatory variables for all SDR cases, and compared with previously reported predictor variables. The AIC is a relative measure of model fit and complexity, with better model quality being indicated by smaller values.^20,21^ Age, gender, diagnosis, and SDR_mean_ were selected as other predictive variables for the variables of interests. Skull thickness, skull area, and SDR_skewness_ were included as known predictor variables for comparison. When candidate variables correlated with the known variables, one was removed to avoid multicollinearity.

### [Statistical Analysis]

To summarize the data, the mean and standard deviation were calculated for normally distributed continuous variables, the median and 25th and 75th percentile values for non-normally distributed continuous variables, and percentages for binary variables. A Q–Q plot evaluated the normality of the data distribution. Group differences were tested using the chi-squared test (for binary variables), Student’s t-test (for normally distributed continuous variables), and the Mann–Whitney U test (for non-normally distributed continuous variables). The significance level was set at p = 0.05. Pearson’s correlation coefficient was used to test correlations. In multiple regression analysis, variables were selected on the basis of clinical judgment, that is, using the basic background information of the patients: age, gender, diagnosis, and SDR_mean_. All statistical analyses were performed with EZR^22^ (Saitama Medical Center, Jichi Medical University, Saitama, Japan), which is a graphical user interface for R (The R Foundation for Statistical Computing, Vienna, Austria). More precisely, it is a modified version of R Commander designed to add statistical functions frequently used in biostatistics.

### [Ethical Consideration]

This study was approved by our institutional ethics committee.

## Results

From April 2020 to March 2023, 171 patients were treated at our institution, 26 of whom had an SDR < 0.40 and were observed 6 months postoperatively with no dropouts. No patient refused to participate in this study, and all underwent thalamotomy for tremor symptoms. Treatment was successful in 15 patients (57.7%). Table 1 shows the patient background characteristics. Patients with essential tremor in the successful treatment group had significantly lower Clinical Rating Scale for Tremor scores than those in the failure group (Table 2).

**Table 1.** Patient background of subcohort with SDR<0.40.

|  | Overall subcohort | failure | success | P |
| --- | --- | --- | --- | --- |
| n | 26 | 11 | 15 |  |
| age | 73.5 [62, 76] | 74 [65, 76] | 74 [65, 76] | 0.146 |
| sex (male) | 18 (69.2%) | 7 (63.6%) | 11 (73.3%) | 0.683 |
| ET | 22 (84.6%) | 9 (81.8%) | 13 (86.7%) | 1 |
| PD | 4 (15.4%) | 2 (18.2%) | 2 (13.3%) | 1 |
| CRST | 35 [16, 40] | 40 [26, 52] | 22 [15, 39] | 0.016† |
| UPDRS* | 14 [13,17] | 18.5 [17, 20] | 12.5 [12, 13] | 0.121 |
| treatment side (left) | 22 (84.6%) | 11 (100%) | 11 (73.3%) | 0.113 |
| SDR<sub>mean</sub> | 0.37 [0.36, 0.39] | 0.37 [0.37, 0.39] | 0.38 [0.37, 0.39] | 0.268 |
| Skull area | 345 [326, 365] | 338 [331, 365] | 351 [324, 362] | 0.659 |
| Skull thickness | 7.40 [6.50, 7.81] | 7.77 [6.29, 8.28] | 7.40 [6.58, 7.78] | 0.938 |
| IA<sub>mean</sub> | 13.3 [12.6, 14.0] | 13.6 [13.2, 14.0] | 13.1 [12.1, 13.6] | 0.103 |

Nominal variable(%), continuous variable, median [25 percentile, 75 percentile], \* UPDRS part II #16 + part III #20-21, maximum 32, Mann-Whitney U test, † P<0.05; ET, essential tremor; PD, Parkinson's disease; CRST, clinical rating scale of tremor; UPDRS, Unified Parkinson's disease rating score; SDR, skull density ratio; IA, incident angle.

**Table 2.** Patient background of full cohort.

|  | full cohort |
| --- | --- |
| n | 171 |
| age | 73 [64, 78] |
| sex (male) | 130 (76.5%) |
| ET | 145 (84.8%) |
| PD | 26 (15.2%) |
| treatment side (left) | 147 (86.0%) |
| SDR<sub>mean</sub> | 0.50 ±0.095 |
| Skull area | 348 ±22.8 |
| Skull thickness | 6.67 [6.12, 7.40] |
| IA<sub>mean</sub> | 13.1 ±0.99 |

Nominal variable (%), normally distributed continuous variable, mean±standard deviation, non-normally distributed continuous variable, median [25 percentile, 75 percentile]; ET, essential tremor; PD, Parkinson's disease; skull density ratio; IA, incident angle.

Univariate analysis of cases with SDR < 0.40 showed that the skull variables with p < 0.05 (not statistically significant because of multiple comparisons) were the mean IA in the D region (IA_D,mean_) and the mean SDR in the G and H regions (SDR_G,mean_ and SDR_H,mean_) (Table 3). IA_D,mean_ tended to be smaller (i.e., closer to vertical) in the successful treatment group, and both SDR_G,mean_ and SDR_H,mean_ also tended to be smaller in this group. Although this may seem paradoxical, it suggested that the mean SDR without G and H (SDR_woGH,mean_) may be relatively larger in the successful treatment group. In fact, SDR_woGH,mean_ tended to be larger in the successful group (Table 3). We also visualized the IA, SDR, and skull thickness in regions A–J in the successful and failure groups, as well as all for cases (Figure 2, Supplementary Tables 1–3).

**Table 3.** Candidates resulting from univariate analysis of subcohort with SDR<0.40.

|  | failure | success | P |
| --- | --- | --- | --- |
| IA<sub>D,mean</sub> | 14.1 [13.4, 15.0] | 11.8 [11.1, 14.2] | 0.012 |
| SDR<sub>G,mean</sub> | 0.36 [0.32, 0.40] | 0.33 [0.30, 0.34] | 0.040 |
| SDR<sub>H,mean</sub> | 0.37 [0.32, 0.45] | 0.31 [0.29, 0.40] | 0.040 |
| SDR<sub>woGH,mean</sub> | 0.37 [0.37, 0.39] | 0.38 [0.37, 0.39] | 0.51 |

median [25 percentile, 75 percentile], Mann-Whitney U test; IA, incident angle; SDR, skull density ratio.

**Figure 2.**
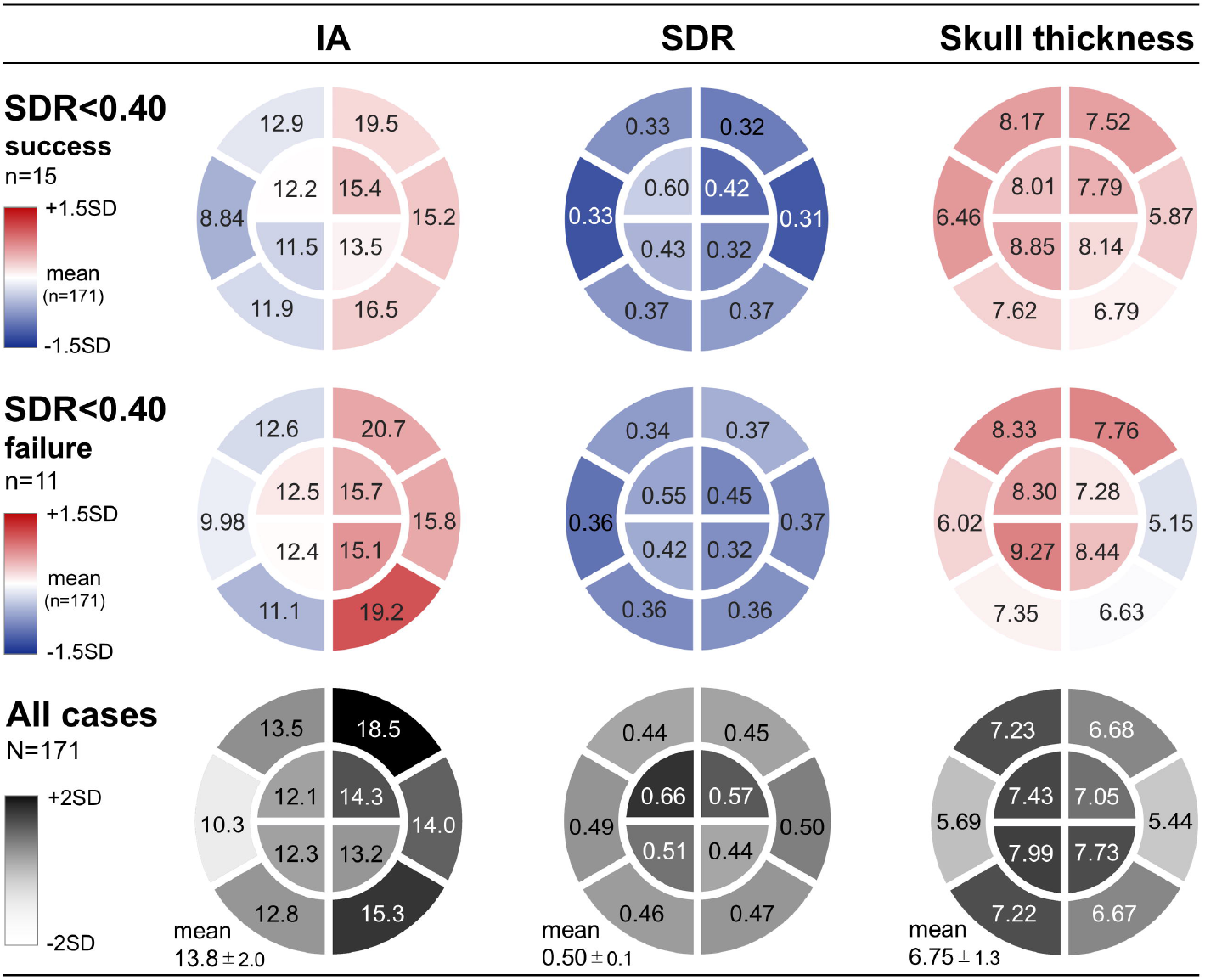
Skull conditions for the treatment success/failure groups and the entire cohort. The upper and middle rows show the difference from the mean of all cases in each section. The lower row shows the difference for each section compared with the mean of the entire skull. IA, incident angle; SDR skull density ratio.

There was a positive correlation between IA_D,mean_ and the mean IA (IA_mean_) (Figure 3A, correlation coefficient: 0.535, 95% confidence interval [CI]: 0.418– 0.634, p = 4.81e-14). No correlation was found between IA_D,mean_ and the ratio of IA_D,mean_ to IA_mean_ (Figure 3B, correlation coefficient: 0.0969, 95% CI: -0.0539– 0.243, p = 0.207). There was a very strong positive correlation between SDR_woGH,mean_ and SDR_mean_ (Figure 4A, correlation coefficient: 0.977, 95% CI: 0.969–0.983, p = 8.4e-116). No correlation was found between the value of SDR_woGH,mean_ and the ratio of SDR_woGH,mean_ to SDR_mean_ (Figure 4B, correlation coefficient: -0.276, 95% CI: -0.409–-0.131, p = 0.000259).

**Figure 3.**
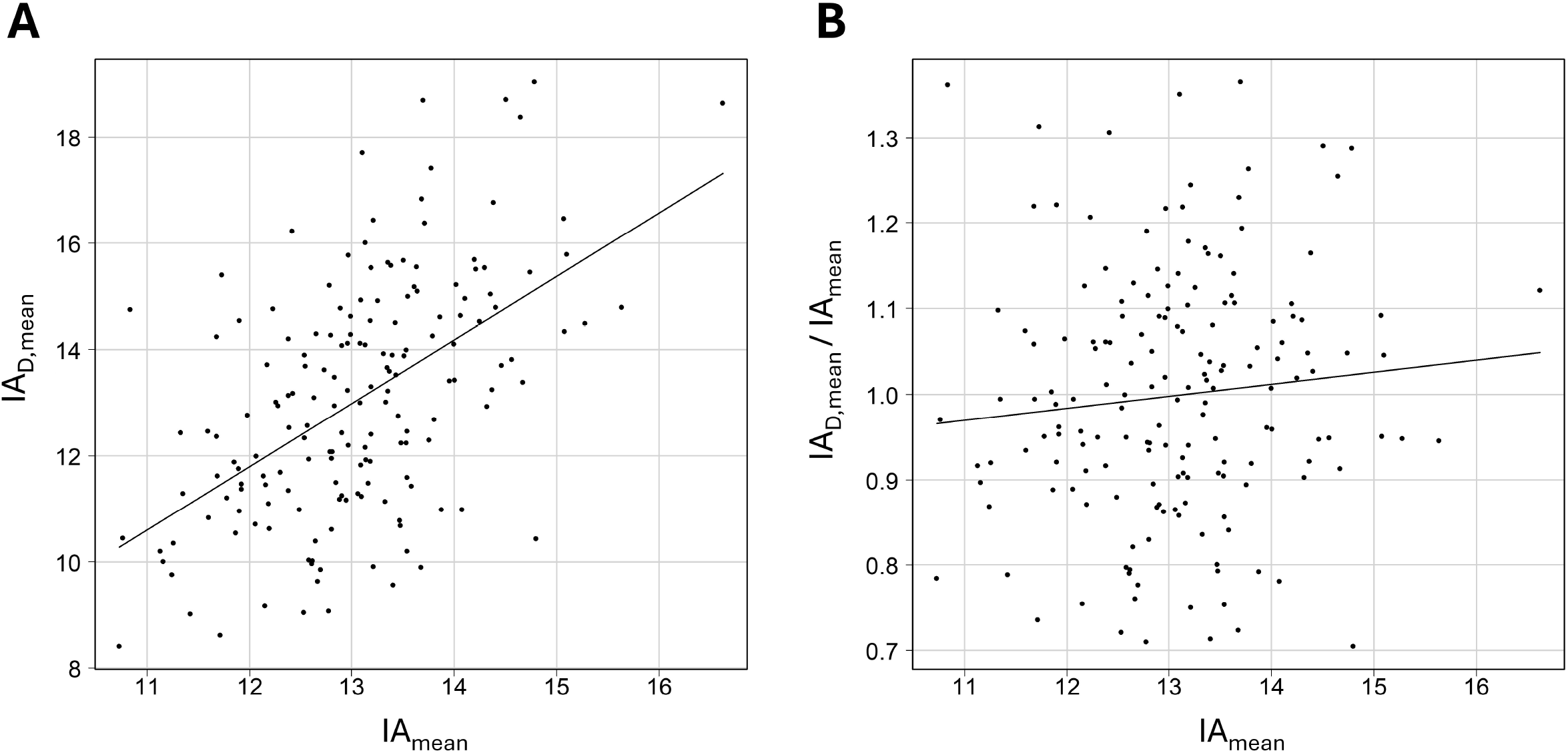
**A**. Correlation between IA_D,mean_ and IA_mean_. Correlation coefficient = 0.535, 95% confidence interval: 0.418–0.634, p = 4.81e-14. IA_mean_, mean incident angle; IA_D,mean_, mean incident angle in the D region. **B**. Correlation between IA_mean_ and the ratio of IA_D,mean_ to IA_mean_. Correlation coefficient = 0.0969, 95% confidence interval: -0.0539–0.243, p = 0.207. IA_mean_, mean incident angle; IA_D,mean_, mean incident angle in the D region.

**Figure 4.**
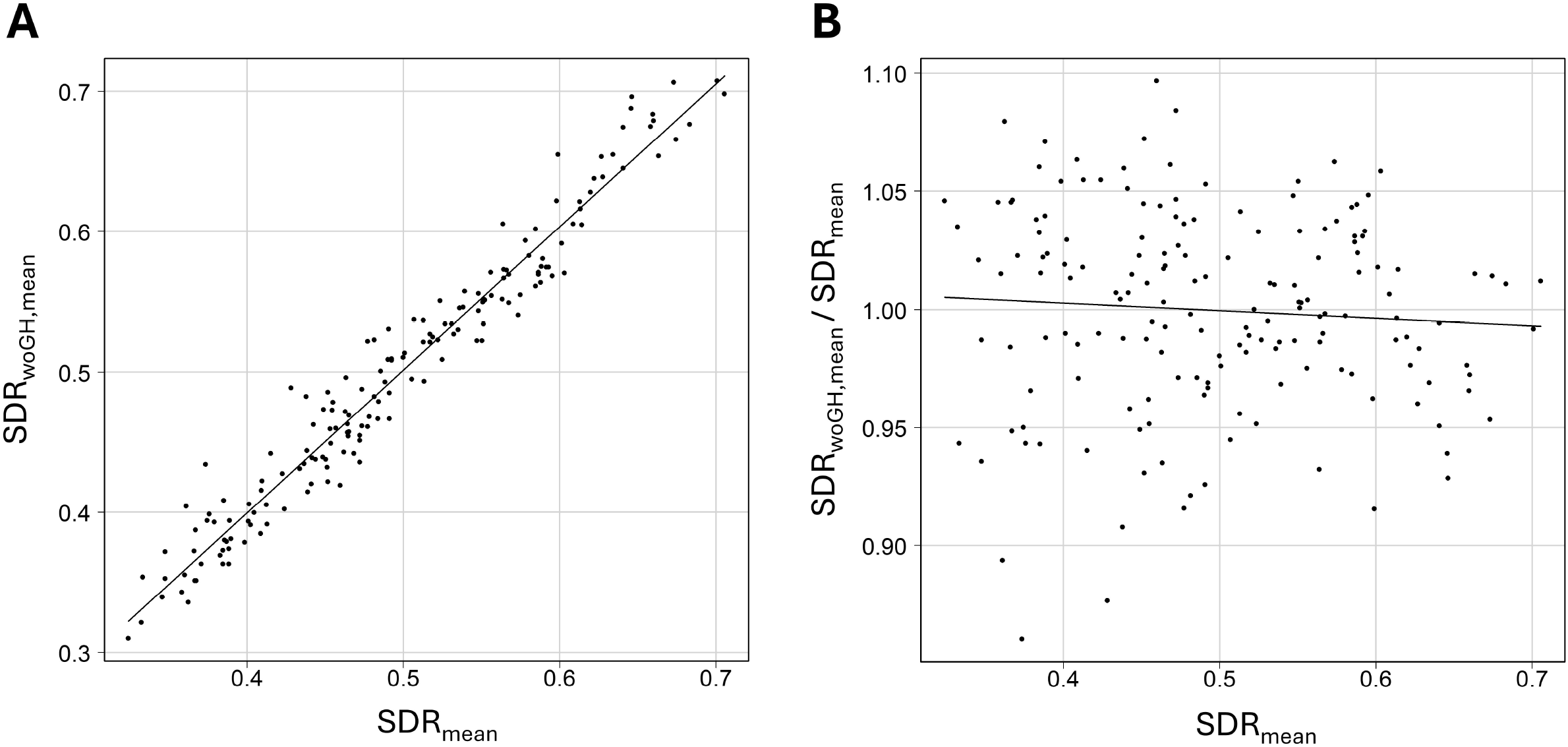
**A**. Correlation between SDR_woGH,mean_ and SDR_mean_. Correlation coefficient = 0.977, 95% confidence interval: 0.969–0.983, p = 8.4e-116. SDR_mean_, mean skull density ratio; SDR_woGH,mean_, mean SDR without G and H. **B**. Correlation between SDR_mean_ and the ratio of SDR_woGH,mean_ to SDR_mean_. Correlation coefficient = -0.276, 95% confidence interval: -0.409– 0.131, p = 0.000259. SDR_mean_, mean skull density ratio; SDR_woGH,mean_, mean SDR without G and H.

On the basis of the results described so far, we compared the predictive performance of IA_D,mean_ and SDR_woGH,mean_ with that of conventional variables using the AIC (Table 4). Compared with the model with an AIC of 777, which was calculated by multiple regression analysis and included maximum temperature as the outcome variable and age, sex, diagnosis, and SDR_mean_ as explanatory variables, the model to which IA_D,mean_ was added had a smaller AIC value (757). This value was also smaller than that calculated by adding IA_mean_ (768), which is the mean of the entire skull, and smaller than that when skull area (776) and skull thickness (778) were added. Similarly, compared with the AIC calculated using age, sex, diagnosis, and SDR_mean_ (777), the AIC calculated using SDR_woGH,mean_ (767) instead of SDR_mean_ was smaller. The AIC calculated using SDR_skewness_ instead of SDR_mean_ was higher (829), suggesting the superiority of SDR_woGH,mean_. Adding IA_D,mean_ further reduced the AIC (750). Our final equation for predicting the maximum temperature rise is as follows (adjusted R^2^: 0.512, F-statistic: 36.5, p < 2.2e-16);

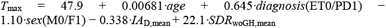

**Table 4.** Akaike information criteria.

| Parameters | $R^2_{\text{adj}}$ | AIC |
| --- | --- | --- |
| age, sex, diagnosis, $\text{SDR}_{\text{mean}}$ | 0.428 | 777 |
| age, sex, diagnosis, $\text{SDR}_{\text{mean}}$, $\text{IA}_{\text{D,mean}}$ | 0.492 | 757 |
| age, sex, diagnosis, $\text{SDR}_{\text{mean}}$, $\text{IA}_{\text{mean}}$ | 0.458 | 768 |
| age, sex, diagnosis, $\text{SDR}_{\text{mean}}$, Skull area | 0.434 | 776 |
| age, sex, diagnosis, $\text{SDR}_{\text{mean}}$, Skull thickness | 0.428 | 778 |
| age, sex, diagnosis, $\text{SDR}_{\text{woGH,mean}}$ | 0.461 | 767 |
| age, sex, diagnosis, $\text{SDR}_{\text{skewness}}$ | 0.221 | 829 |
| age, sex, diagnosis, $\text{SDR}_{\text{woGH,mean}}$, $\text{IA}_{\text{D,mean}}$ | 0.512 | 750 |

AIC, Akaike information criteria, $\text{SDR}_{\text{woGH}}$ , $R^2_{\text{adj}}$ , adjusted R squared.

### [Representative Case]

Left thalamotomy for essential tremor refractory to drug treatment in a male in his 60’s with CRST 22, SDR_mean_ 0.38, Skull area 315, and skull thickness 7.4mm. SDR_woGH,mean_ was 0.41 and IA_D,mean_ was 14.2°. In the treatment stage, the temperature rise was 54 °C at 20000 J (1000 W·21 sec), 54 °C at 30000 J (1200 W·26 sec), and 54 °C at 36000 J (1200 W·21 sec). The operative record noted that the temperature rise was similar to that of a high SDR_mean_. Postoperatively, the CRST improved to 6, and was still CRST 6 at 6 months postoperatively, indicating that the therapeutic effect was maintained.

## Discussion

In this study, we explored variables predicting treatment success or failure in low-SDR cases using our institution’s cohort of patients with SDR < 0.40 and the data of the total cohort. Treatment success was achieved in 57% of thalamotomies with SDR < 0.40. Univariate analysis of the cohort with SDR < 0.40 identified IA_D,mean_, SDR_G,mean_, and SDR_H,mean_ as candidate predictors. In a multiple regression model predicting maximum temperature rise using the entire cohort, IA_D,mean_ and SDR_woGH,mean_ showed better AICs than the conventional variables. These results indicate that IA_D,mean_ and SDR_woGH,mean_ are generally significant variables for predicting temperature rise in MRgFUS and may also predict treatment efficacy in low-SDR cases.

Among the results related to SDR, SDR_G,mean_ and SDR_H,mean_, i.e., bilateral temporal SDRs, showed small p-values in univariate analysis when SDR < 0.40, and both SDRs were lower in the successful treatment group. Although larger SDRs are generally considered more effective for sonication,^1-7^ our result appears contrary to this. Because the analysis was limited to cases with SDR < 0.40, we interpreted the lower bilateral temporal SDRs in the successful group as indicating that SDR_woGH,mean_ was higher in this group. Indeed, the SDR_woGH,mean_ tended to be larger in the successful group within the SDR < 0.40 cohort. Across all cohorts, the multiple regression model using SDR_woGH,mean_ showed better AIC than that using SDR_mean_, suggesting that SDR_woGH,mean_ is a more effective predictor of maximum temperature. Additionally, SDR_woGH,mean_ was correlated with SDR_mean_, but the ratio of SDR_woGH,mean_ to SDR_mean_ was not. This means that SDR_woGH,mean_ can vary even within low SDR_mean_ cases, suggesting that a good treatment effect can be expected even with a low SDR_mean_ if SDR_woGH,mean_ is relatively favorable.

Basic studies on ex vivo skulls may provide some interpretation of these results. In a study by Riis TS et al.^23^ measuring the ultrasound transmission of three ex vivo skulls, the bilateral temporal regions were markedly more efficient in ultrasound transmission and had less phase distortion than other skull regions. In other words, the bilateral temporal regions had a significant advantage in FUS. Combined with our findings, this suggests that the bilateral temporal regions transmit ultrasound so effectively that poor SDR may not impact treatment success.

Another notable result of this study was that, in the cohort with SDR < 0.40, the successful treatment group tended to have a smaller IA_D,mean_, i.e., a more vertical ultrasound IA, in the posterior half of the parietal region on the treated side, than the failure group. Furthermore, a multiple regression model predicting the maximum temperature across all cohorts showed a better AIC with IA_D,mean_ than with IA_mean_. Although it is generally believed that ultrasound has less loss to reach intracranial areas when the entrance angle to the skull is closer to vertical,^1,11,14,24^ the IA to the skull is determined by the skull’s three-dimensional shape and size, target coordinates and eccentricity, and position relative to the head frame.^14,25^ Moreover, the skull is non-spherical and complex.^14,25^ Our findings suggest that extracting the posterior half of the parietal region on the treatment side may be more effective in predicting treatment outcome in terms of IA than averaging over the entire skull. In general, averaging the whole does not always provide a good representation of the actual overall performance, possibly leading to the “fallacy of averages”.^26,27^ The IA of individual ultrasound elements is influenced by many complex stereospatial factors, and IA_mean_ may discard valuable information through averaging. IA_D,mean_ will differ even in cases with the same IA_mean_, and our findings suggest that IA_D,mean_ may be a more appropriate predictor of treatment success than IA_mean_.

In our study, skull thickness did not show as much predictive ability as SDR_woGH,mean_ and IA_D,mean_. Basic studies have shown that as skull thickness increases, ultrasound transmission decreases^28^, energy attenuation increases^23^, and phase distortion increases^23^. However, some clinical studies have reported an association with the temperature rise^14^ and required energy^9^, whereas others found no association with temperature^59^ or heating efficiency^29^. There are likely still gaps in our understanding of the diagnostic value of skull thickness that need further investigation.

### Limitations

This exploratory study has several limitations. Because of the limited number of cases with SDR < 0.40, multivariate validation was performed using the data of all SDR cases. Also, selection bias may occur when enrolling low-SDR patients into treatment. Additionally, the outcome variable was the maximum temperature rise, not treatment success or failure. Further validation is needed to confirm whether the same results can be replicated in patients with low SDRs alone.

## Conclusions

We explored the skull conditions that differentiate treatment success from failure in low-SDR cases. The ultrasound angle of incidence in the parietal region on the treatment side, as well as SDR calculated excluding bilateral temporal areas, emerged as candidates. These two variables predicted temperature rise more effectively than conventional variables across the entire cohort, irrespective of the SDR.

## Data Availability

All data produced in the present study are available upon reasonable request to the authors

## Acknowledgments

Thank individuals who contributed to the study or manuscript preparation but who do not fulfill all the criteria of authorship.

