## Supplementary Tables for "The Potential Role of the Regional Skull Conditions in Predicting the Efficacy of Transcranial Magnetic Resonance-guided Focused Ultrasound in Patient with Low Skull Density Ratio"

**Supplementary Table 1** Skull conditions of all cases (n=171). IA, incident angle; SDR skull density ratio.

|  | IA | SDR | Skull thickness |
| --- | --- | --- | --- |
| mean | 13.8 ±2.00 | 0.50 ±0.10 | 6.75 ±1.30 |
| A | 12.14 ±2.53 | 0.66 ±0.17 | 7.43 ±1.49 |
| B | 14.32 ±2.95 | 0.57 ±0.14 | 7.05 ±1.51 |
| C | 12.33 ±2.61 | 0.51 ±0.15 | 7.99 ±1.68 |
| D | 13.26 ±2.82 | 0.44 ±0.15 | 7.73 ±1.78 |
| E | 13.50 ±3.40 | 0.44 ±0.14 | 7.23 ±1.58 |
| F | 18.53 ±4.01 | 0.45 ±0.14 | 6.68 ±1.45 |
| G | 10.32 ±2.65 | 0.49 ±0.13 | 5.69 ±1.22 |
| H | 14.01 ±3.39 | 0.50 ±0.16 | 5.44 ±1.36 |
| I | 12.81 ±3.28 | 0.46 ±0.12 | 7.22 ±1.44 |
| J | 15.38 ±3.65 | 0.47 ±0.14 | 6.67 ±1.34 |

**Supplementary Table 2** Skull conditions of treatment success group (n=15). Median [minimum, maximum]. IA, incident angle; SDR skull density ratio.

|  | IA | SDR | Skull thickness |
| --- | --- | --- | --- |
| A | 12.2 [9.18, 15.4] | 0.60 [0.48, 0.72] | 8.01 [6.09, 9.03] |
| B | 15.4 [11.4, 19.0] | 0.42 [0.29, 0.51] | 7.79 [5.42, 11.0] |
| C | 11.5 [9.48, 15.3] | 0.43 [0.33, 0.51] | 8.85 [6.98, 9.66] |
| D | 13.5 [7.78, 15.8] | 0.32 [0.20, 0.41] | 8.14 [5.41, 12.2] |
| E | 12.9 [8.24, 15.8] | 0.33 [0.26, 0.41] | 8.17 [6.30, 10.9] |
| F | 19.5 [15.9, 25.6] | 0.32 [0.26, 0.37] | 7.52 [6.03, 9.70] |
| G | 8.84 [7.12, 14.6] | 0.33 [0.27, 0.40] | 6.46 [4.46, 10.6] |
| H | 15.2 [12.3, 17.4] | 0.31 [0.24, 0.50] | 5.87 [4.00, 9.55] |
| I | 11.9 [7.97, 19.2] | 0.37 [0.29, 0.53] | 7.62 [6.49, 10.2] |
| J | 16.5 [12.0, 19.6] | 0.37 [0.34, 0.45] | 6.79 [5.65, 9.72] |

**Supplementary Table 3** Skull conditions of treatment failure group (n=11). Median [minimum, maximum]. IA, incident angle; SDR skull density ratio.

|  | IA | SDR | Skull thickness |
| --- | --- | --- | --- |
| A | 12.5 [8.73, 13.7] | 0.55 [0.44, 0.76] | 8.30 [6.14, 10.5] |
| B | 15.7 [12.7, 17.6] | 0.45 [0.37, 0.57] | 7.28 [5.02, 9.74] |
| C | 12.4 [8.94, 15.7] | 0.42 [0.31, 0.50] | 9.27 [7.17, 11.7] |
| D | 15.1 [13.2, 18.7] | 0.32 [0.11, 0.49] | 8.44 [6.36, 12.8] |
| E | 12.6 [10.7, 13.9] | 0.34 [0.26, 0.40] | 8.33 [5.38, 10.0] |
| F | 20.7 [18.0, 24.1] | 0.37 [0.24, 0.48] | 7.76 [4.65, 8.90] |
| G | 9.98 [8.10, 12.9] | 0.36 [0.31, 0.43] | 6.02 [4.58, 9.05] |
| H | 15.8 [12.1, 17.4] | 0.37 [0.30, 0.53] | 5.15 [4.11, 8.75] |
| I | 11.1 [10.3, 14.7] | 0.36 [0.27, 0.49] | 7.35 [6.27, 10.4] |
| J | 19.2 [12.1, 20.9] | 0.36 [0.29, 0.46] | 6.63 [5.63, 10.3] |
